# Mortality among Brazilian male police officers and firefighters over three decades

**DOI:** 10.64898/2026.10.08.26365007

**Authors:** Elaine Mélo Teixeira, Fernanda Camargo Damacena, Fernando Barbosa de Souza, Robson Dettmann Jarske, Luiz Guilherme Grossi-Porto, Eliana Zandonade, Karla Nívea Sampaio

## Abstract

**Background:** Long-term mortality data among Brazilian police officers and firefighters are scarce. This study investigated the leading causes of death in Brazilian male military police officers and firefighters over three decades, comparing their mortality and mean age at death with those of the male population.

**Methods:** We retrospectively analyzed death records of male military police officers and firefighters from Espírito Santo (ES), Brazil, from 1988 to 2019. Mortality was analyzed across three decades and the standardized mortality ratios (SMRs) for the three leading causes were calculated using the male populations of ES and Brazil as references.

**Results:** Overall, 2,198 deaths were identified, with a mean age at death of 59 ± 18.5 years, compared with 60.5 ± 23.9 years among men in ES and 63.4 ± 23.7 years among Brazilian men. Mean age at death was 36 ± 9 years among active-duty and 68 ± 13 years among inactive personnel. Across the study population, the leading causes of death were circulatory system diseases (32%), external causes (26%) and neoplasms (15%). Circulatory disease mortality was higher in the second and third decades compared with both reference populations, whereas external-cause mortality shifted from higher, in the first two decades, to lower than expected in the last decade. Neoplasm mortality was higher relative to Brazil only in the last decade.

**Conclusion:** Mortality patterns differed by cause and over time. The younger mean age at death among active-duty personnel, together with cause-specific excess mortality, highlights the importance of long-term occupational health surveillance and preventive strategies.

## 1. Introduction

Work-related stress has been associated with an increased risk of adverse health outcomes, as workers may be exposed to both physical and psychological stressors [1]. Likewise, frequent occupational exposure to hazardous or toxic substances may adversely affect health [2,3]. Some professionals are more susceptible to occupational stress and unhealthy conditions due to the nature of their work. Among these, firefighters and police officers stand out due to various physically and psychologically demanding activities and occupational hazards, such as heat and smoke exposure, shift work, work overload, exposure to violence, and physical exhaustion, making these professionals more susceptible to psychological distress, emotional exhaustion, and adverse health outcomes [1,4,5].

Some studies have pointed to lower longevity among these professionals than in the general population, potentially associated with occupational exposures and their health effects [6–9], although findings varied across different countries [10,11]. Exposure to various carcinogenic substances during firefighting or frequent exposure to airborne pollutants during police traffic surveillance may contribute to an increased risk of cancer incidence and mortality among these workers [2,10]. Stressful and physically demanding activities, such as firefighting and pursuing suspects, have been associated with an increased risk of sudden cardiac death [3,9]. Exposure to violence and traumatic events is also inherent to these occupations and may contribute to an increased risk of death by external causes, including occupational injuries, violence and suicide [12].

In a previous study, we reported a high prevalence of obesity and overweight, important cardiovascular risk factors, in a population of firefighters from Southeastern Brazil [13]. Reduced cardiorespiratory fitness and a high prevalence of overweight and obesity have also been reported among firefighters from the Federal District of Brazil [14]. Similarly, a recent study reported a high prevalence of chronic diseases and cardiovascular risk factors among military police officers and firefighters in Southern Brazil [15]. In a population of firefighters from the Southeast of Brazil, we recently identified that nearly 50% (45.6%) presented elevated blood pressure levels, including both pre-hypertension and hypertension [16]. This condition was significantly associated with other critical cardiovascular risk factors, including central obesity, hyperglycemia, and hypertriglyceridemia[16]. Despite evidence of a substantial burden of cardiovascular risk factors among Brazilian police officers and firefighters, which may contribute to adverse long-term health outcomes [9,17,18], information on causes of death and mortality among these occupational groups in Brazil remains scarce.

Thus, our study investigated the main causes of death over three decades among a population of military Brazilian male police officers and firefighters and compared their mortality and mean age at death with those of the general male population. We hypothesized that mortality from cardiovascular diseases among these workers would be higher than expected based on the general male population.

## 2. Methods

### 2.1 Study Design

A retrospective analytical study was carried out using secondary data from death certificates of police officers and firefighters from the state of Espírito Santo (ES), a state located in Southeastern Brazil. The study period extended from 1988 to 2019, allowing the construction of a time series and analyses across three decades. The analysis was restricted to deaths occurring up to 2019 to characterize the pre-pandemic mortality profile of police officers and firefighters. Deaths occurring from 2020 onwards were not included because the COVID-19 pandemic introduced a distinct epidemiological period, with potential direct and indirect effects on cause-specific mortality and death certification, which could bias comparisons across the three historical decades analyzed.

### 2.2 Study Population

The study population comprised military firefighters and police officers from the state of Espírito Santo (ES) who died between 1988 and 2019. The data were collected from death certificates and other documents attached to the pension payment *post-mortem* process filed at their military pension institution. Data collection at the Pension Fund Institution occurred between January 2018 and December 2019, and an active search for the latest records continued until October 2022. In ES state, the Fire Brigade remained part of the Military Police until 1997, when it became a separate institution. Until that date, officers who performed duties as firefighters were officially registered as police officers. For that reason, the two categories of workers were treated together in the present study.

To compare mortality among officers with mortality in the general populational (see statistical analysis for description), mortality data for the populations of ES state and Brazil were obtained from the Brazilian Mortality Information System (*Sistema de Informações sobre Mortalidade*; SIM), available through the Ministry of Health’s Department of Informatics of the Unified Health System (DATASUS). Data on deaths among males were selected by residence and age group, using the International Classification of Diseases 9^th^ Revision (ICD-9) for death data between 1988 and 1995 and the 10^th^ Revision (ICD-10) for data between 1996 and 2019. The resident population for the studied period was obtained from the demographic statistics available on the DATASUS website. Population data from 1988 to 2012 were obtained from the 2010 Brazilian Census and intercensal estimates, and data from 2013 and 2019 were obtained from the population projections available on the DATASUS website. Since the minimum age for joining the military service is 18 years old, population data collection began with the age group 15-19 years old. The number of professionals in service per year was provided by the Human Resources Department of the Military Police and Fire Brigade of the ES state.

### 2.3 Inclusion and Exclusion Criteria

All deaths of male firefighters and police officers that occurred between 1988 and 2019 were included. Only male deaths were included in the present study due to the small number of female deaths (N=14). Death certificates for which the date of birth was not available were excluded from the study (N=3).

### 2.4 Study Variables

Data collected were the date of birth, date of death, cause of death, employment status (active-duty or inactive) and skin color. The age-at-death variable was determined by subtracting the date of birth from the date of death. The variable skin color was categorized as white, mixed, and black, according to the information provided in the death certificates.

The underlying cause of death was considered the main cause of death. The causes of death described on the certificates were categorized with the help of a pathologist, following the chapters of the 9^th^ and 10^th^ revisions of the ICD.

### 2.5 Statistical Analyses

Categorical variables were presented as absolute and relative frequency and quantitative data as mean ± standard deviation. Categorical data were analyzed by chi-square test. The Prais-Winsten regression model was used to analyze the temporal trend in the annual number of deaths from 1988 to 2019. The annual number of deaths was log-transformed and used as the dependent variable, while the year of death was used as the independent variable. Based on the b1 coefficient estimated from the regression analysis, the Annual Percent Change (APC) and its respective 95% Confidence Interval (CI) were calculated [19]. An increased trend is considered when the APC is positive, a decreased trend when it is negative, and an unchanged trend when the APC is not statistically different from zero (p>0.05).

Mean age at death among military personnel was calculated directly from the study database and separately for active-duty and inactive personnel. For comparison, mean age at death in the general male populations of ES and Brazil was estimated for the same study period (1988– 2019) using mortality data obtained from SIM/DATASUS. Deaths were grouped by age, and mean age at death was calculated as a weighted mean using the midpoint of each age interval and the corresponding number of deaths. For the open-ended category of ≥80 years, an upper boundary of 120 years was assumed, corresponding to a midpoint of 100 years. The standard deviation was calculated using the corresponding age-group frequencies.

Deaths were further analyzed by decades, considering the intervals 1990-1999, 2000-2009 and 2010-2019, and excluding the years 1988 and 1989 from these analyses. For each year from 1990 to 2019, the military population was determined by considering individuals who had already joined the institution and were alive in that year and was stratified by age group (15– 19, 20–29, 30–39, 40–49, 50–59, 60–69, 70–79, and ≥80 years). Standardized Mortality Ratios (SMRs) were estimated for the three leading causes of death by decade using indirect age standardization, as described in Gordis [20]. Briefly, age-specific mortality rates were calculated for the male populations of ES and Brazil by dividing the number of deaths in each age group by the corresponding population and multiplying by 100,000. These age-specific mortality rates were then applied to the corresponding age groups of the military population to estimate the number of expected deaths. Expected deaths were summed for each decade, and the SMR was calculated as the ratio between the total number of observed and expected deaths for each of the three main causes of death. The 95% Confidence Interval for the SMR was calculated according to Vandenbroucke [21]. Analyses were performed using Microsoft Excel 365 and Statistical Package for Social Sciences (SPSS, version 25, USA) software.

### 2.6. Ethical Aspects

The study was approved by the Military Fire Service (CBMES) and the Military Police Service (PMES) of the ES state and by the Human Research Ethics Committee of the Federal University of Espírito Santo (CEP-UFES), under approval number; CAAE: 70725817.1.0000.5060.

## 3. Results

### 3.1. Deaths by age and year of occurrence

Over the 32 years, 2,198 deaths were recorded among male police officers and firefighters in the state of Espírito Santo, after excluding female deaths and records without date of birth. The minimum age at death was 19 and the maximum was 108 years old, with a mean age at death of 59 years (SD = 18.5). The mean age at death was 36 years old (SD = 9) among active-duty military personnel and 68 years old (SD = 13) among inactive personnel. The mean age at death among men in the Brazilian general population was 63.4 years (SD = 23.7) and 60.5 (SD = 23.9) years among men in ES during the same study period.

The annual number of deaths from 1988 onwards is shown in Figure 1. The overall average number of deaths per year was 69 officers (SD = 12.2). The Prais-Winsten regression showed a significant increasing trend in the annual number of deaths over the study period, with an Annual Percentage Change (APC) of 0.92% (CI: 0.49% to 1.36%; p=0.01).

**Figure 1.**
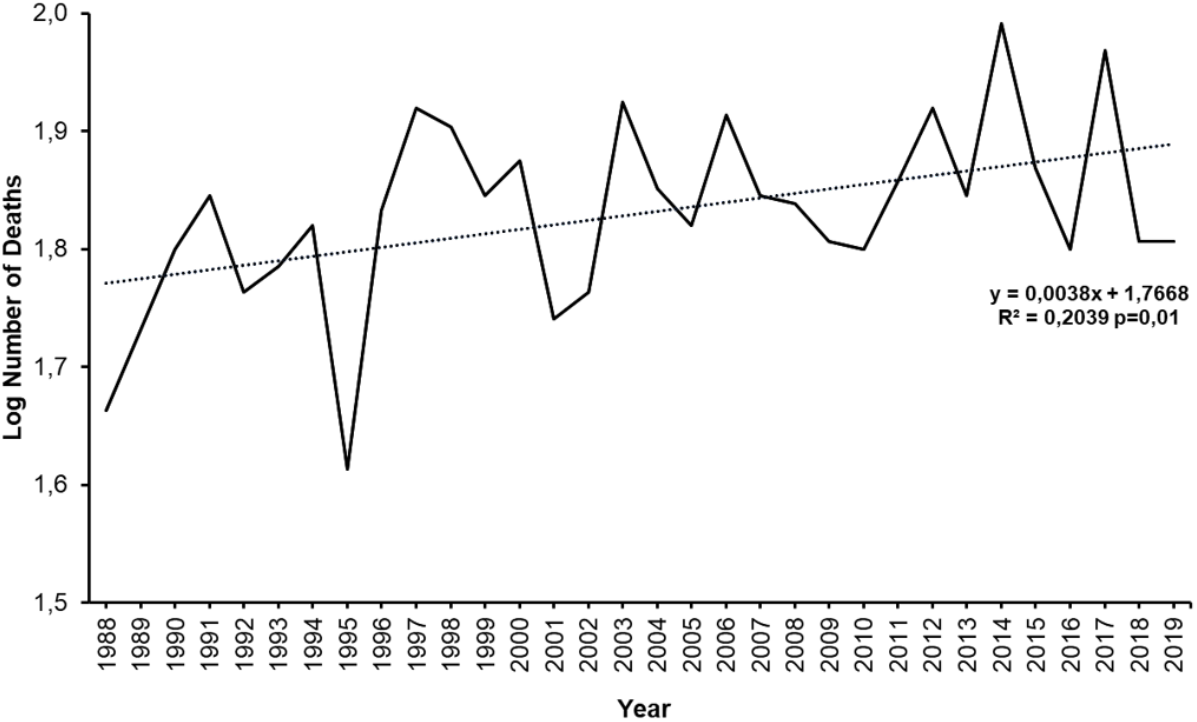
Deaths by year of occurrence among police officers and firefighters from Espírito Santo state between 1988 and 2019. Temporal trend in the annual number of deaths analyzed by Prais-Winsten regression using log-transformed values.

### 3.2. Causes of death

The causes of death are depicted in Figure 2. The leading causes of death among police officers and firefighters were diseases of the circulatory system (32%), external causes (26%) and neoplasms (15%). Deaths from undetermined causes accounted for 7% of all deaths.

**Figure 2.**
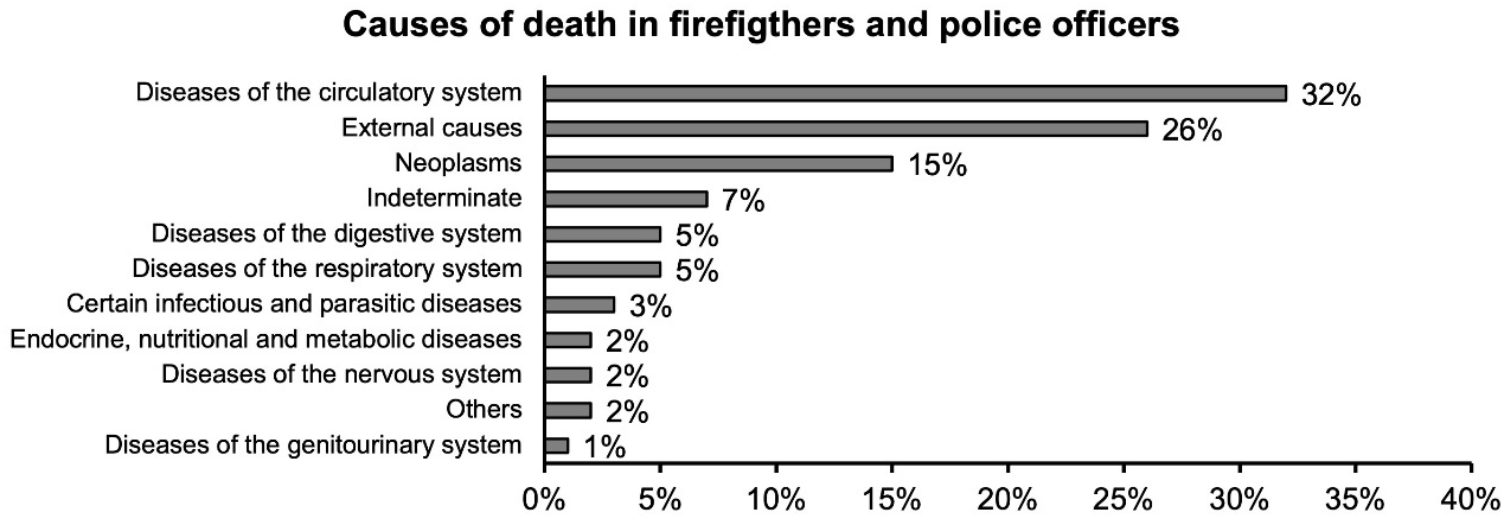
Causes of death among police officers and firefighters from Espírito Santo state between 1988 and 2019.

### 3.3. Deaths by decade

The number of deaths from all causes in the first decade (1990 to 1999) was 660, which corresponds to 31% of deaths occurring between 1990 and 2019. From 2000 to 2009, the number of deaths increased to 695 (33%), and in the last decade, total deaths encompassed 743 officers (36%). Higher ages at death were observed in subsequent decades for the three leading causes of death (Figure 3). The analysis also revealed outliers for neoplasm deaths among young officers in the first decade and for external causes in older workers in the first and second decades.

**Figure 3.**
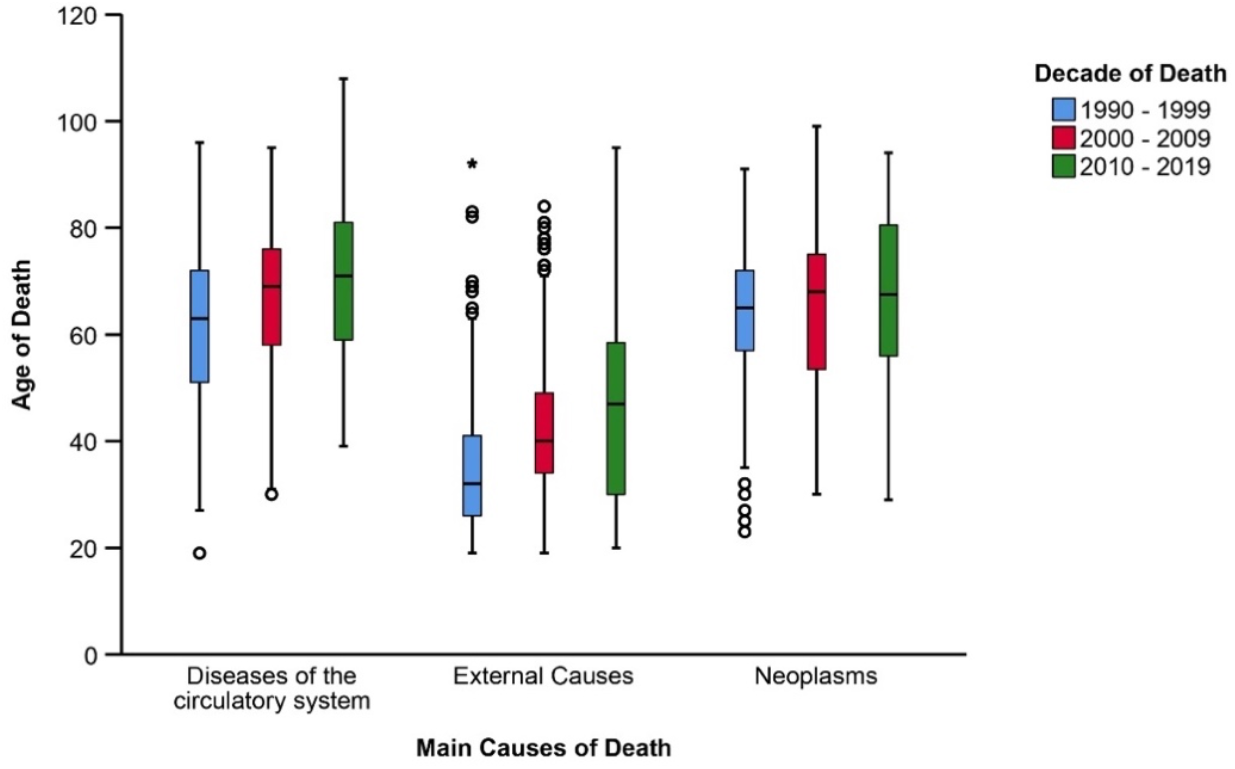
Box plot showing the distribution of age at death by decade for the three main causes of death among police officers and firefighters of Espírito Santo state. Circles represent outliers and asterisks represent extreme outliers, according to the interquartile range criterion.

### 3.4. Main causes of death *vs* skin color

Pearson’s chi-squared test showed a statistically significant association between the three main causes of death and skin color (p<0.01). Among white military police officers and firefighters who died from one of the three main causes, diseases of the circulatory system accounted for 42.2% (N=147) of deaths, followed by neoplasms at 29.3% (N=102) and external causes at 28.4% (N=99). Similar percentages were observed among mixed skin color police officers and firefighters, with 48.8% (N=303) of deaths from diseases of the circulatory system, 28.0% (N=174) from external causes and 23.2% (N=144) from neoplasms. In contrast, among black police officers and firefighters, 64.9% (N=37) of deaths from the three main causes were due to diseases of the circulatory system, with 17.5% (N=10) each from external causes and neoplasms.

### 3.5. Mortality from the main causes in police officers and firefighters compared with the populations of ES and Brazil

Table 1 shows the Standardized Mortality Ratios (SMRs) for the three leading causes (diseases of the circulatory system, external causes, and neoplasms) in police officers and firefighters, using the male population of ES state and Brazil as reference populations.

**Table 1.** Standardized Mortality Ratios (SMRs) for the three main causes of death among police officers and firefighters, using the male populations of Espírito Santo state and Brazil as reference populations, across three decades.

| Cause | Decade* | Standardized Mortality Ratios (SMRs) for Espírito Santo |  |  |  | Standardized Mortality Ratios (SMRs) for Brazil |  |  |  |
| --- | --- | --- | --- | --- | --- | --- | --- | --- | --- |
|  |  | Expected deaths | Observed deaths | SMR | 95% CI | Expected deaths | Observed deaths | SMR | 95% CI |
| Diseases of the circulatory system | 1 | 183.7 | 197.0 | 1.07 | 0.92 to 1.23 | 173.2 | 197.0 | 1.14 | 0.98 to 1.31 |
|  | 2 | 195.8 | 226.0 | 1.15 | 1.01 to 1.31 | 168.7 | 226.0 | 1.39 | 1.17 to 1.52 |
|  | 3 | 168.4 | 251.0 | 1.49 | 1.31 to 1.68 | 164.5 | 251.0 | 1.53 | 1.34 to 1.72 |
| External Causes | 1 | 129.6 | 237.0 | 1.83 | 1.60 to 2.07 | 97.4 | 237.0 | 2.43 | 2.13 to 2.76 |
|  | 2 | 149.9 | 188.0 | 1.25 | 1.08 to 1.44 | 105.3 | 188.0 | 1.79 | 1.53 to 2.06 |
|  | 3 | 224.6 | 116.0 | 0.52 | 0.43 to 0.62 | 147.8 | 116.0 | 0.78 | 0.65 to 0.94 |
| Neoplasms | 1 | 69.4 | 77.0 | 1.11 | 0.87 to 1.38 | 70.1 | 77.0 | 1.10 | 0.86 to 1.36 |
|  | 2 | 91.4 | 96.0 | 1.05 | 0.85 to 1.28 | 84.4 | 96.0 | 1.14 | 0.92 to 1.38 |
|  | 3 | 156.4 | 156.0 | 1.00 | 0.84 to 1.16 | 95.1 | 156.0 | 1.64 | 1.39 to 1.91 |

\*Decade 1: 1990 to 1999; Decade 2: 2000 to 2009; Decade 3: 2010 to 2019.

Police officers and firefighters had a higher than expected mortality from diseases of the circulatory system in the second and third decades compared with both reference populations. In the first decade, the SMRs was not significantly different from 1.0 for either ES or Brazil.

Mortality from external causes was higher than expected in the first two decades compared with both reference populations and lower than expected in the third decade.

Neoplasm mortality was not significantly different from expected mortality based on the ES population in any decade. Compared with the Brazilian population, neoplasm mortality was higher than expected only in the third decade.

## 4. Discussion

This study provides a long-term assessment of cause-specific mortality among police officers and firefighters from ES, Brazil, using state and national populations as references. Our study found a mean age at death of 59 years among male police officers and firefighters from ES, compared with 60.5 years among men in ES and 63.4 years among men in Brazil, with a particularly low mean age at death of 36 years among active-duty personnel. An increasing trend in the annual number of deaths over the years, with diseases of the circulatory system, external causes, and neoplasms being the three leading causes of death. Police officers and firefighters had higher than expected mortality from diseases of the circulatory system in the second and third decades and from external causes in the first two decades, compared both with reference populations. In the third decade, mortality from external causes was lower than expected relative to both populations, whereas neoplasms mortality was higher than expected only relative to the Brazilian population.

The mean age at death found for police officers and firefighters in our study was 36.0 years old among active personnel (SD=9) and 68.0 years (SD=13) among inactive personnel. In American firefighters, the mean age at death from work-related cardiac fatalities was 49 years old (SD=9) [22]. A similar age at death has been observed among American police officers, with most occupational deaths occurring between 40 and 60 years of age [18]. The mean age at death among active-duty personnel in our study was lower than the ages reported in these studies. However, direct comparisons are limited because the cited studies evaluated occupational fatalities, whereas our study included deaths from all causes among active-duty personnel, regardless of whether they occurred on duty or not. The differences observed may be associated with occupational profiles, variations in population characteristics and methodological criteria. The overall mean age at death in our study was 59.0 years, compared with 60.5 years among men in ES and 63.4 years among men in Brazil during the same study period. These descriptive differences do not establish a lower life expectancy among police officers and firefighters, given the different age structures of the populations. Occupational exposure to physical and psychosocial stressors, together with the high prevalence of cardiometabolic risk factors reported in these professions, may contribute to adverse health outcomes [1,3,4,13–16,18,23].

We observed an increasing trend in the annual number of deaths, with the highest number in the last decade, corresponding to 36% of deaths recorded between 1990 and 2019. Higher ages at death were observed in subsequent decades for the three leading causes of death. A study investigating firefighter fatalities in the United States over two decades, 1990-2000 and 2002-2012, also showed an increase in the number and age at death from cardiovascular events over time [24]. Among American police officers, an increase in the number of deaths related to occupational activities was also observed between 1997 and 2018 [18]. The increase in the annual number of deaths observed in our study may reflect changes in the size of the military workforce, as well as factors related to occupational health and length of service. However, an increase in mortality risk over time cannot be inferred from the analysis of absolute numbers of deaths alone. The contribution of the natural aging of the cohort to the increase in deaths over time also cannot be discarded, particularly deaths related to neoplasms and diseases of the circulatory system. Similarly, the higher ages at death observed in subsequent decades may partly reflect changes in the age structure of the workforce rather than necessarily indicating increased longevity. The possible impact of changes in working conditions throughout this long period of analysis is beyond the scope of our study, but the potential effects of health promotion and wellness programs implemented over this period cannot be disregarded.

Diseases of the circulatory system were the leading cause of death followed by external causes and neoplasms among police officers and firefighters from our study. In this respect, our findings are consistent with those of previous studies conducted in different countries. The SMR showed higher than expected mortality from diseases of the circulatory system in the last two decades compared with both reference populations, and from neoplasms in the last decade compared with the general Brazilian population. Many studies have also reported circulatory system diseases and neoplasms as the leading causes of death among firefighters and police officers from different countries [9,11,24–28].

Between 1950 and 2005, white male police officers from Buffalo, New York, showed a higher standardized mortality ratio for all diseases of the circulatory system (SMR=1.11; 95% CI: 1.02-1.19) when compared with the US general population [26]. More recent data from the same cohort, but including a broader period (1950-2018), reported mortality from circulatory system diseases that was not significantly different from that expected based on the general population (SMR=0.99; 0.92-1.06) [11]. On the other hand, the mortality from malignant neoplasms was higher than expected in both study periods (1950-2005; SMR=1.32; 95%CI 1.19-1.46 and 1950-2018: SMR=1.14; 95%CI 1.04-1.25), although the SMR was lower in the study with the extended follow-up period [11,26].

Among firefighters, different mortality patterns have been reported depending on the population studied, time interval, and the cause of death. In a cohort of American firefighters from San Francisco, Chicago, and Philadelphia, between 1950 and 2016, mortality from ischemic heart disease was not significantly different from that expected in the general population (SMR = 0.98; 95% CI: 0.95–1.01), whereas mortality from cancer was higher than expected (SMR = 1.12; 95% CI: 1.08–1.16) [17]. Differences between the SMRs reported in different studies may be related to population differences, study periods, and temporal stratification, as illustrated by the differences observed across decades in our study. The excess cancer mortality reported in the American cohort is consistent with the higher than expected neoplasm mortality observed in the last decade of our study when Brazil was used as the reference population. In that decade, the SMR indicated higher than expected neoplasm mortality among military personnel compared with the Brazilian general population, but not with the population of ES. One possible explanation for this finding is differences in the higher completeness of death registration in the Mortality Information System (SIM) in ES relative to many other Brazilian states, which may have affected the estimated mortality rates [29].

Several well-known risk factors for cardiovascular diseases and neoplasms, such as stressful and life-threatening activities [11], shift work with changes in sleep patterns [5,30], insufficient physical activity, poor diet, overweight, and obesity [4,13–15], hypertension [31], excessive alcohol consumption, smoking, and a sedentary lifestyle [2,32], are common in this occupational group. In addition, some activities such as firefighting involve intense physical effort and exposure to toxic compounds, including cyanide, carbon monoxide [33] and carcinogens (benzene, polycyclic aromatic hydrocarbons, and formaldehyde) [27], often released in high quantities during the combustion process in fires. Similarly, frequent exposure to airborne carcinogens from motor vehicle exhaust can also be associated with an increased risk of cancer incidence and mortality among traffic police officers [2]. Exposure to particulate matter, for instance, can reduce the efficiency of DNA repair and increase the frequency of mutagenesis [34].

External causes were the second leading cause of death among our population of Brazilian male firefighters and police officers. The SMR for external causes was significantly higher than expected in the first two decades compared with both reference populations, but mortality was significantly lower than expected in the last decade. External causes have also been reported as one of the main causes of death in other populations of firefighters and police officers, but unlike our findings in the first two decades, some studies have reported lower than expected mortality from external causes in these professionals when compared with the general population [10,11,26,28].

In our study, the high mortality from external causes among firefighters and police officers in the first two decades, particularly among younger officers, may partly reflect the historical context of violence in Brazil [35], especially among professionals who deal with crime and insecurity on a daily basis. Police officers deal constantly with violence in their work routines. Combat situations in military operations, added to other factors such as work overload and inadequate and obsolete equipment, expose police officers to health-threatening risks, particularly those carrying out patrolling and crime-fighting activities [12]. Constant exposure to violence and traumatic events may also contribute to psychological distress and suicidal behavior among police officers [12,36]. Suicide has also been identified as an important concern among firefighters [37]. In addition, adverse working conditions and the risk of physical injury may also contribute to deaths from external causes among firefighters and police officers [1,3,11,18].

In contrast, mortality from external causes was significantly lower than expected relative to both reference populations in the last decade, a finding that may involve different factors. This reduction may be associated with a change in the age structure of the group, including a possible reduction in the proportion of younger professionals as the workforce aged, which could also contribute to the higher age at death observed in the subsequent decades. Added to this, particularly in firefighters, the emergence of more modern and widely available respiratory protection equipment [38] may contribute to reducing the inhalation of toxic substances during fires as well as other related fatalities. In fact, among American firefighters, a reduction in fatalities over time led a significant decline in overall deaths between two periods [39].

Importantly, some initiatives may have contributed to improving the health of these professionals over the study period. In 1992, a Military Police Hospital was created and in 1997, the first Preventive Health Actions Program began to develop preventative and educational activities among this occupational group [40]. In 2017, the Military Fire Brigade of the ES created and implemented the project “Bom Estar” (“Wellness Project”), which aimed to conduct a health assessment of these workers as a strategy to improve their health conditions [41]. Although the effects of this initiative on mortality could not be assessed within the study period, such programs may contribute to improving the long-term health of these professionals [41]. Of note, advances in health technologies concerning diagnosis, treatment, and prevention, as well as changes in the behavior and lifestyle of the population [42], which includes police officers and firefighters [43], may also have influenced mortality patterns over time. The healthy worker effect should also be considered when comparing these professionals with the general population, as health-based selection is part of their admission criteria and may contribute to lower mortality in this occupational group [10]. Nevertheless, the excess mortality observed for some causes and periods suggests that occupational risk factors warrant further investigation.

We must recognize some limitations of the present study. Sociodemographic and health information on the professionals included in the study was not available, which could help explain some of the present findings. In addition, apart from occupational exposure, factors such as lifestyle habits, length of career, activities performed as a police officer or firefighter, and history of pre-existing illnesses could be related to the deaths observed, but these data were not available. Also, the inability to discriminate between the two occupational groups limits our ability to identify differences in mortality between police officers and firefighters, although the currently available evidence points to adverse health outcomes in both groups. The analysis of the temporal trend was based on absolute numbers of deaths and therefore did not account for changes in the size of the military workforce. In addition, comparisons of mean age at death with the general population were descriptive and did not account for differences in population age structure. Despite these limitations, the analysis of 2,198 deaths over 32 years provides substantial long-term evidence on mortality among police officers and firefighters from ES, Brazil, a population for which studies of this scope remain scarce. Although our results are limited to one Brazilian state, our findings are likely generalizable to other similar populations, given that, in Brazil, firefighters and police officers acting in patrolling service are military personnel with similar job organization and job-related activities nationwide. The comparison of cause-specific mortality across three decades using both state and national reference populations further strengthens the contribution of our study.

## 5. Conclusion

Our results show a particularly low mean age at death among active-duty firefighters and police officers and a high proportion of deaths from diseases of the circulatory system, external causes, and neoplasms. Mortality from diseases of the circulatory system was higher than expected in the last two decades compared with both reference populations. In the last decade, mortality from external causes was lower than expected relative to both populations, whereas neoplasm mortality was higher than expected only relative to the Brazilian population. These findings highlight the need for preventative and surveillance actions and the importance of health monitoring among these professionals.

## 8. Authors contributions

EMT and FCD collected the data; EMT, FCD, FBS, EZ, and KNS analyzed and interpreted the data; EMT wrote the first draft of the manuscript; RDJ helped with death categorization; LGP contributed to the interpretation of the results and critically revised the manuscript; KNS supervised EMT, conceived the study, and revised the final version of the manuscript. All authors read and approved the final version of the manuscript.

## 9. Data availability

The data that support the findings of this study are available from the corresponding author upon reasonable request and subject to approval by the relevant institutions

## 10. Preprint statement

This manuscript is a preprint and has not undergone peer review.

## 11. Use of generative AI

ChatGPT (OpenAI) was used solely for English-language editing and to improve clarity and readability. All suggested edits were reviewed and approved by the authors, who take full responsibility for the final content.

